# Cost-effectiveness analysis of patent foramen ovale closure versus medical therapy alone after cryptogenic stroke

**DOI:** 10.1101/2021.10.13.21263767

**Authors:** Yoko Shijoh, Shota Saito, Zhehao Dai, Sachiko Ohde

**Author notes:** Corresponding author (YS).

## Abstract

**Background:** Closure of a patent foramen ovale reduces the risk of recurrent stroke compared with medical therapy alone in young patients with cryptogenic strokes revealed by randomized control trials. Some cost-effectiveness analyses outside Japan have shown that patent foramen ovale closure is cost-effective, but no studies have examined cost-effectiveness in Japan. The objective of this study is to assess cost-effectiveness, from the perspective of a Japanese healthcare payer, of patent foramen ovale closure versus medical therapy alone for the patients with patent foramen ovale related cryptogenic strokes.

**Methods:** A cost-effectiveness study was conducted by developing a decision tree and a Markov model. Probabilities and a 5.9-year time horizon followed the RESPECT study. Utilities and costs were based upon published studies and assumptions. The model cycle was one month. All assumptions were assessed by experts, including a cardiologist and a statistical expert. The target population comprised patients with cryptogenic stroke and patent foramen ovale, aged 60 years or younger. Incremental cost-effectiveness ratio was evaluated. Then one-way sensitivity analyses and probabilistic sensitivity analyses were conducted to assess robustness.

**Results:** Incremental cost-effectiveness ratio of patent foramen ovale closure compared with medical therapy was estimated at ¥3,318,152 per quality-adjusted life years gained. One-way sensitivity analysis showed that the stable state utility score difference between patent foramen ovale closure and medical therapy had the largest impact on incremental cost-effectiveness ratio. Patent foramen ovale closure is cost-effective at a stable state utility score difference of >0.051, compared with medical therapy. Probabilistic sensitivity analyses demonstrated that patent foramen ovale closure was 50.3% cost-effective with a willingness-to-pay threshold of ¥5,000,000 / quality-adjusted life years.

**Conclusions:** From a healthcare payer perspective, patent foramen ovale closure may be cost-effective compared with medical therapy for Japanese patients with cryptogenic stroke who were ≤60 years.

## Introduction

Patent foramen ovale (PFO) is an opening in the septum between the right and left atria that failed to close at birth. Relative to the total number of general autopsy findings, the prevalence of PFO is reportedly 26% (1). Under certain hemodynamic conditions, such a PFO can be forced open by a pressure gradient that favors right-to-left shunting, thereby enabling blood and bloodborne substances to pass from the venous to the arterial circulation (2). This is the mechanism whereby PFO is associated with paradoxical embolism, which is the most common cause of cryptogenic stroke in young adults (3).

Approximately 25% of cerebral infarctions are of unknown cause, but some are thought to be due to thrombi from the right heart system that entered the left heart system through right-left shunts, such as a PFO. These are called paradoxical embolisms. However, thrombi in the right heart system are often not detected in cases of cerebral infarction with PFO, and no treatment showing clear effectiveness has been established for prevention of recurrent stroke in cases in which aspirin is commonly used (4). Possible treatments to prevent recurrent stroke in cryptogenic stroke patients with a PFO include medical treatment with antiplatelet agents or anticoagulants, percutaneous PFO closure, and surgical PFO closure.

Recently, several randomized controlled trials showed that percutaneous transcatheter closure reduces the risk of recurrent stroke compared with medical therapy alone, among relatively young patients with cryptogenic stroke complicated by a PFO (5–7). The Japanese government approved a percutaneous transcatheter closure device, AMPLATZER PFO Occluder, and its procedure in 2019 for secondary prevention of ischemic stroke in patients ≤60 years who had a cryptogenic stroke that was probably attributable to a PFO. Until 2020, AMPLATZER PFO Occluder was the only device approved in Japan for PFO closure. Soon after the government approved the device and procedure, the Japan Stroke Society, the Japanese Circulation Society, and the Japan Cardiovascular Intervention Treatment Society released guidelines for percutaneous PFO closure for cryptogenic stroke (4). These guidelines were based upon evidence from three randomized control trials (RCTs) in 2017: RESPECT (5), REDUCE (6) and CLOSE (7), which focused on efficacy and safety with an appropriate patient background. The guidelines raised the clinical issue of postoperative atrial fibrillation rate, which was higher in the PFO closure group than in a non-closure group in a meta-analysis that included the three aforementioned RCTs (8–10).

Reduction of the recurrent stroke rate is important, not only from a clinical perspective, but also from a health economic perspective. Some cost-effectiveness analyses outside Japan have compared PFO closure and medical therapy alone for cryptogenic stroke patients and have shown that PFO closure is cost-effective for quality-adjusted life years (QALYs) gained. (11–16). However, there have been no cost-effectiveness analyses in Japan and such analyses, which include cost information, utility scores, and a lifetable would be more convincing than cost-effectiveness analyses done outside Japan. Therefore, our objective was to assess the cost-effectiveness, from a Japanese healthcare payer perspective, of PFO closure with AMPLATZER PFO Occluder for risk reduction of recurrent stroke in patients with cryptogenic stroke that was probably attributable to a PFO, compared to medical therapy alone.

## Method

### Model overview

We developed a decision tree and Markov model to assess the cost-effectiveness of PFO closure compared with medical therapy alone. (Fig 1) For the Markov model, we proposed a model consisting of four health states: stable after cryptogenic stroke, post-minor recurrent stroke, post-moderate recurrent stroke, and death. (Fig 1) We used a modified Rankin Scale (mRS) to categorize post-minor stroke as mRS 0 to 2 and post-moderate recurrent stroke as mRS 3 to 5. The time horizon was derived from the RESPECT study median follow-up period, which was 5.9 years (5). The model cycle was one month. QOL scores for each condition can be calculated, and to calculate an incremental cost-effectiveness ratio (ICER), the incremental cost increase per PFO closure was divided by the QALYs gained by the closure. Five million yen per QALY was determined as the cost-effectiveness willingness-to-pay that the Japanese government sets as the ICER threshold in the cost-effectiveness evaluation system. We conducted one-way sensitivity analyses with varied key assumptions to assess robustness. The model was discounted at 2.0%, which is a basic discount rate in the guidelines for preparing cost-effectiveness evaluations for the central social insurance medical council in Japan. All analyses were performed using TreeAge Pro software.

**Fig 1.**
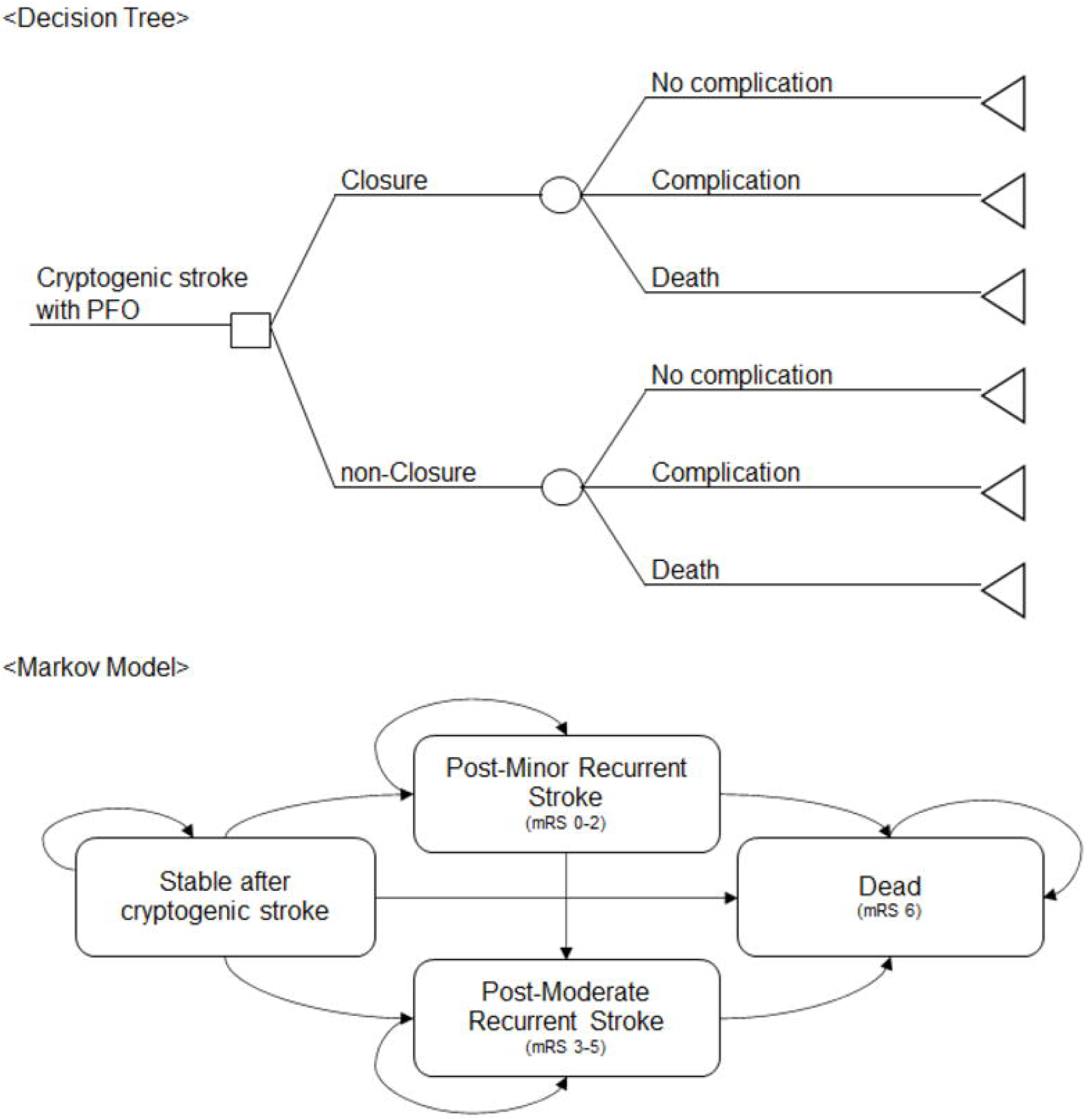
The model structure. A decision tree (upper picture) shows that a patient who had a PFO enters this model after a cryptogenic stroke and is assigned either a PFO closure or medical treatment. PFO closure has three possible outcomes: complication, non-complication, or death. The Markov model (lower picture) shows four health states: stable after cryptogenic stroke, post-minor recurrent stroke, post-moderate recurrent stroke, and death. We used a modified Rankin Scale to categorize strokes. Abbreviations. PFO, patent foramen ovale; mRS, modified Rankin Scale.

### Patients

The target population included cryptogenic stroke patients who had a PFO, aged ≤60 years, based on the indication for AMPLATZER™ PFO Occluder in Japan. The Japanese government approved AMPLATZER™ PFO Occluder based on the RESPECT study results of “rates of 0.58 events per 100 patient-years and 1.07 events per 100 patient-years, respectively (hazard ratio with PFO closure vs. medical therapy, 0.55; 95% confidence interval [CI], 0.31 to 0.999; p=0.046 by the log-rank test).” Thus, patients in the model represent the RESPECT trial, a multi-center, randomized, open-label, controlled clinical trial with blinded adjudication of end-point events (5).

### Probabilities and Utilities

Probabilities for each outcome obtained from published literature are shown in Table 1. Probabilities for recurrent ischemic stroke were estimated on a monthly cycle based on the RESPECT study primary endpoint. We assumed that the recurrent ischemic stroke rate persists during the simulation. In the decision tree, we set atrial fibrillation and flutter as procedural complications, because calculation of low-incidence, serious adverse events (SAEs) is impractical and should only address clinically problematic SAEs. This conclusion was based on guidance from the Japan Stroke Society, the Japanese Circulation Society, and the Japan Cardiovascular Intervention Treatment Society for percutaneous patent foramen ovale closure for latent cerebral infarction. This statement argued that meta-analysis clarified the effectiveness of this treatment, despite the increased risk of postoperative atrial fibrillation (8–10).

**Table 1.** Probabilities of events, life years, and utility scores for base model inputs and sensitivity analyses.

|  | Base value |  | Reference |
| --- | --- | --- | --- |
|  | Closure | Medical therapy |  |
| Procedural / Device deaths | 0.00% | — | RESPECT study (5) |
| Recurrent clinical ischemic stroke | 0.58% | 1.07% | RESPECT study (5) |
| Atrial fibrillation or flutter | 1.40% | 0.83% | RESPECT study (5) |
| Stable state utility | 0.88 | 0.8 | Tirschwell et al. (15) |
| Annual probability of natural death |  |  | MHLW life table 2018 (26) |

|  | Base value |  | Range / Distribution |  | Reference |
| --- | --- | --- | --- | --- | --- |
|  |  | lower | Upper | Distribution |  |
| Time horizon | 5.9 | 5.9 | 38.3 | - | RESPECT study (5), WHO Life expectancy (27) |
| Closure risk ratio of recurrent stroke | 0.54 | 0.31 | 0.999 | Triangular | RESPECT study (5) |
| PFO closure atrial fibrillation or flutter HR | 0.50 | 1.69 | 10.12 | Triangular | REDUCE study (6) |
| Post-stroke mortality rate within 1 year | 6.00% | 5.40% | 6.60% | - | Linxin Li et al. (20) |
| Post-stroke mortality rate within 5 years | 24.9% | 22.4% | 27.4% | - | Linxin Li et al. (20) |
| Post-stroke mortality rate within 10 years | 45.5% | 41.0% | 50.1% | - | Linxin Li et al. (20) |
| % of ischemic stroke mRS0-2 | 68.0% | 61.2% | 74.8% | - | Grau AJ et al. (21) |
| % of ischemic stroke mRS3-5 | 32.0% | 28.8% | 35.2% | - | Grau AJ et al. (21) |
| Medical therapy stable state utility | 0.800 | 0.720 | 0.880 | - | Tirschwell et al. (15) |
| Stable state utility difference | 0.080 | 0.000 | 0.080 | Triangular | expert opinion |
| Minor stroke utility | 0.779 | 0.701 | 0.857 | - | Hattori et al. (23) |
| Moderate stroke utility | 0.338 | 0.304 | 0.372 | - | Hattori et al. (23) |
| Atrial fibrillation utility | 0.725 | 0.653 | 0.798 | - | Reynolds MR et al. (22) |
| Medical device and procedural costs | ¥1,830,000 | ¥1,464,000 | ¥2,196,000 | gamma | expert opinion |
| Clinical moderate stroke | ¥1,758,423 | ¥1,406,738 | ¥2,110,108 | - | Kamae et al. (25) |
| Clinical minor stroke | ¥1,024,306 | ¥819,445 | ¥1,229,167 | - | Kamae et al. (25) |
| Aspirin | ¥9,408 | ¥7,526 | ¥11,290 | - | Claims data. |
| DOAC after atrial fibrillation | ¥185,196 | ¥148,157 | ¥222,235 | - | Claims data. |
| Follow-up cost first year in closure | ¥56,556 | ¥45,245 | ¥67,867 | - | Claims data. |
| Follow-up cost after 2-year in closure | ¥39,960 | ¥31,968 | ¥47,952 | - | Claims data. |
| Follow-up cost non-closure | ¥31,164 | ¥24,931 | ¥37,397 | - | Claims data. |
| Post-clinical moderate stroke cost | ¥4,135,524 | ¥3,308,419 | ¥4,962,629 | - | Hattori et al. (23) |
| Post-clinical minor stroke cost | ¥1,940,172 | ¥1,552,138 | ¥2,328,206 | - | Hattori et al. (23) |

Notes. Probabilities, utility scores, and cost ranges were set at $\pm 10\%$ , $\pm 10\%$ and $\pm 20\%$ , respectively, based on expert opinion.
Abbreviations. MHLW, ministry of health, labour and welfare; PFO, patent foramen ovale; HR, hazard ratio; mRS, modified Rankin Scale; DOAC, novel direct oral anticoagulants.

**Table 2.** Base case cost-effectiveness analysis.

| Treatment | Total cost | Incremental Cost | Total QALY | Incremental Effectiveness | ICER |
| --- | --- | --- | --- | --- | --- |
| Medical Therapy | ¥ 836,280 |  | 4.385 |  |  |
| Closure | ¥ 2,377,004 | ¥ 1,540,724 | 4.849 | 0.464 | 3318152 |

Abbreviations. QALY, quality-adjusted life-years; ICER, incremental cost-effectiveness ratio.

We assumed that the utility score of each health status remained unchanged until the health status changed. The previous assumption of a stable utility score was used from the cost-effectiveness analysis, which was 0.88 for closure-stable utility and 0.8 for medical therapy-stable utility (15). This assumption was based on expert opinion and some previous studies showing a positive impact of the closure procedure on quality of life. Mirzada N (17) explained that the prospect of effective secondary prevention of ischemic recurrences could certainly contribute to better physical, mental, and social functioning in patients who undergo PFO closure. Results reported by Evola S (18) indicated a reasonable association with improvement of migraines, due to a reduction in the frequency and severity of migraine attacks. Lelakowska M (19) also reported that the utility score value of the SF-36 total score was markedly higher 6 months after PFO closure compared with pre-PFO closure.

Based on previous mortality rate reports on cryptogenic stroke patients (20), we used average values of mortality indices categorized into three spans: post-stroke mortality within 1 year, post-stroke mortality within 5 years, and post-stroke mortality within 10 years. Recurrent clinical ischemic stroke was assumed to be 68% for post-minor strokes and 32% for post-moderate strokes, assumptions supported by Grau AJ (21).

We set atrial fibrillation utility using a previous study (22), and minor stroke utility and moderate utility using a Japanese study calculated with the modified Rankin Scale and EQ-5D Japanese version (23).

### Cost and Lifetime table

Our analysis was conducted from a healthcare payer perspective using direct medical costs, including costs for PFO closure procedures, closure complications, medications, follow-up, and stroke care. Procedure costs, medication costs, and follow-up costs were derived from Japanese claim data (24). Treatment costs for acute clinical ischemic stroke and post-clinical ischemic stroke were obtained from previous studies in Japan ^23, 25^. We used the 2018 Japanese lifetime table to obtain the annual probability of natural death (26).

### Sensitivity analysis

One-way sensitivity and probabilistic sensitivity analyses evaluated the impact of assumptions shown by the sensitivity analysis range in Table 1. Lifetime horizon was set as the upper value of time to follow-up, which was 38.3 years, calculated as 84.2 (average life expectancy in Japan) minus 45.9 (mean age of patients in the RESPECT study (5)). We set 5.9 years as the lower value of the time horizon, which is the base case value derived from the RESPECT study (5) median follow-up period. The PFO closure risk ratio of recurrent stroke was obtained from the RESPECT study (5) and we used 95% confidence intervals of the PFO closure risk ratio of recurrent stroke (0.31-0.999) as the lower and upper range. In the RESPECT study (5), the rate of atrial fibrillation or flutter did not differ significantly between the PFO closure and medical therapy groups. The risk ratio was 1.69 and 95% confidence interval was 0.50 to 5.73. We use 1.69 for the base value and 0.50 for the lower value. However, the meta-analysis found that “the rate of newly detected atrial fibrillation in the PFO closure plus medical therapy group and the medical therapy alone group were 4.3% and 0.7%, respectively, which shows that PFO closure plus medical therapy significantly increased the risk of newly detected atrial fibrillation by more than 4 times compared with medical therapy alone (RR 4.69, 95% CI 2.17 to 10.12)”(8). We thus used the upper 95% confidence interval limits of newly detected atrial fibrillation in the meta-analysis as the upper range of atrial fibrillation or flutter rate hazard ratio between closure and non-closure. Other probability scores were set at ±10%, based on expert opinion.

Previous cost-effectiveness studies have shown that stable state utility scores between closure and medical therapy have the widest range for ICERs (11,13–15). No studies report a difference in stable utility scores between closure and medical therapy, and stable utility scores were thus all based on assumption. To clarify the threshold of a stable utility score difference for cost-effectiveness, we set a lower stable utility score difference of 0 and an upper stable utility score difference of 0.08, which was the baseline. Other utility scores and costs were set at ±10% and ±20%, respectively, based on expert opinion.

Cost-effectiveness acceptability curves were determined for the probabilistic sensitivity analysis on the parameter uncertainty at a 5.9-year time horizon. Triangular or gamma distributions were chosen for each parameter (Table 1) and the model was run 10000 times.

## Results

### Base case analysis

According to the analysis results, PFO closure yielded 4.849 QALYs at a cost of ¥2,377,004 and medical therapy yielded 4.385 QALYs at a cost of ¥836,280 over a 5.9-year time horizon (Table. 2). ICER of PFO closure compared with medical therapy was estimated at ¥3,318,152 per QALY gained; therefore, PFO closure is cost-effective compared with medical therapy, based on the ICER threshold set by the Japanese government in the cost-effectiveness evaluation system.

### Sensitivity analysis

Results of one-way sensitivity analysis indicated that the stable state utility score difference between closure and medical therapy had the largest impact on the ICER (Fig 2). With a lower value estimate of 0.00, ICER in the sensitivity analysis was ¥46,327,317 per QALY gained, exceeding acceptable limits of cost-effectiveness. To be cost-effective, thresholds of the stable state utility score difference between closure and medical therapy should be 0.051 (Fig 3).

**Figure 2.**
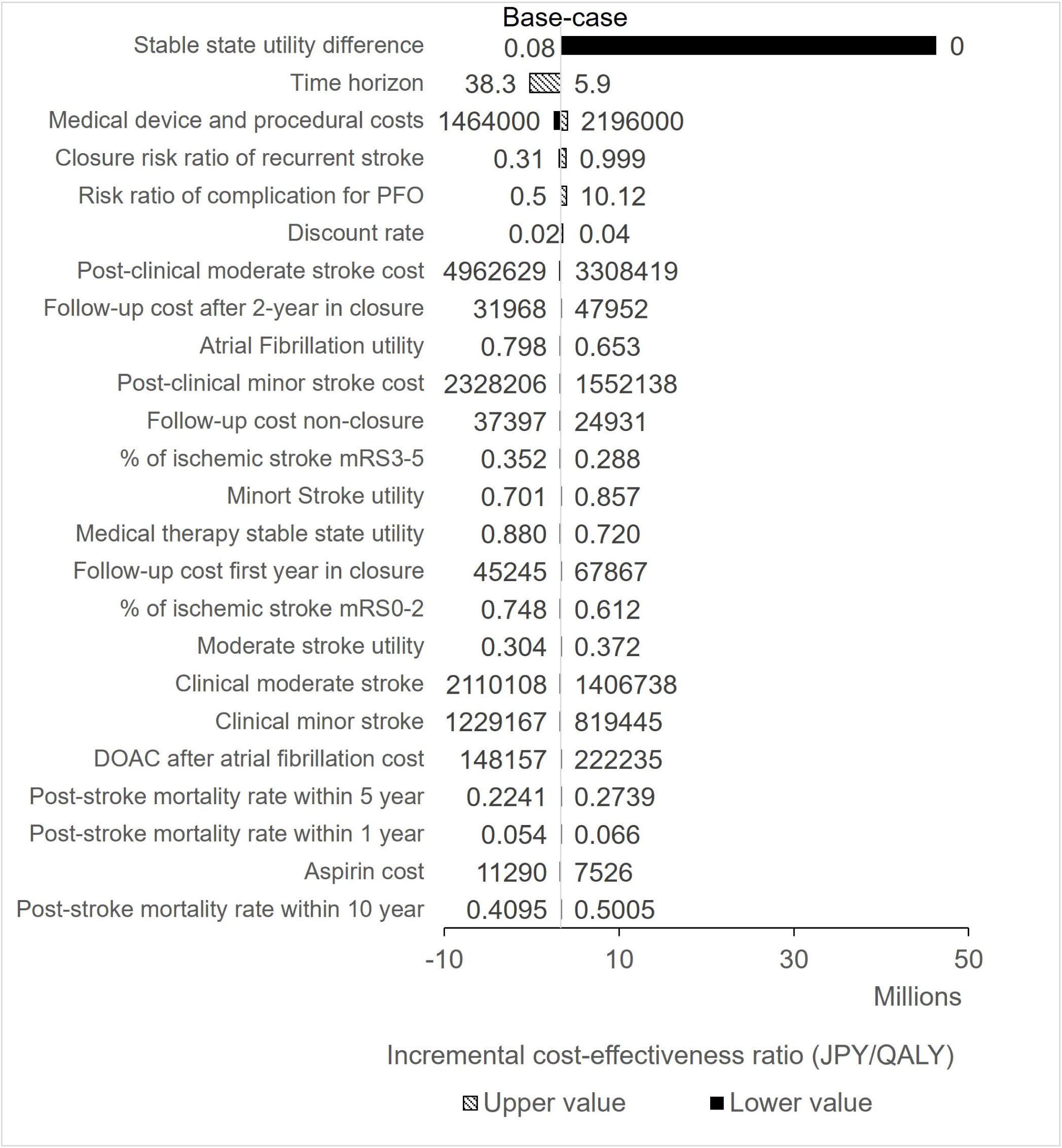
One-way sensitivity analysis of PFO closure VS medical therapy. The impact of parameter variation using a willingness-to-pay threshold of ¥5,000,000/QALY. The hatched bars and black bars show the upper (high estimate) and lower values of the parameter, respectively. Bars are aligned in order of impact from largest to smallest. Abbreviations. PFO, patent foramen ovale; mRS, modified Rankin Scale; DOAC, novel direct oral anticoagulants.

**Figure 3.**
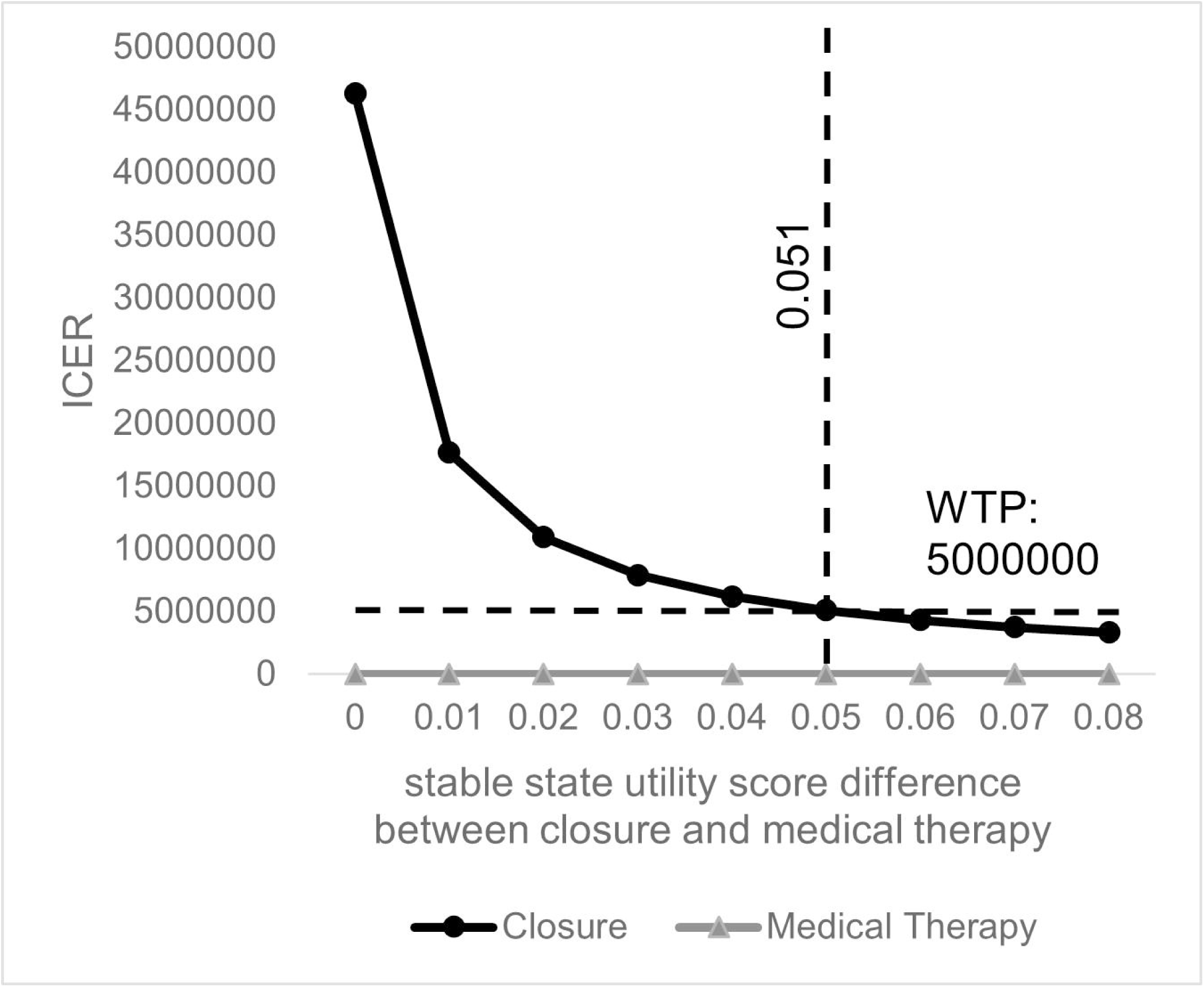
One-way sensitivity analysis: The stable state utility score difference between PFO closure and medical therapy. The stable state utility score difference between PFO closure and medical therapy, which is the largest impact of parameter variation, is shown with ICER. The dashed line represents a willingness-to-pay (WTP) threshold of ¥5,000,000/QALY and a utility score difference of 0.051 at that WTP threshold. Abbreviations. WTP, willingness-to-pay; ICER, incremental cost-effectiveness ratio.

The second largest impact on ICER was the time horizon. PFO closure at the lifetime horizon, which was 38.3 years setting as the upper value of time to follow-up, changed to a dominant economic strategy, resulting in cost reduction and gain in QALY with a base case value having a stable state utility score difference compared with medical therapy (Figure 4).

**Figure 4.**
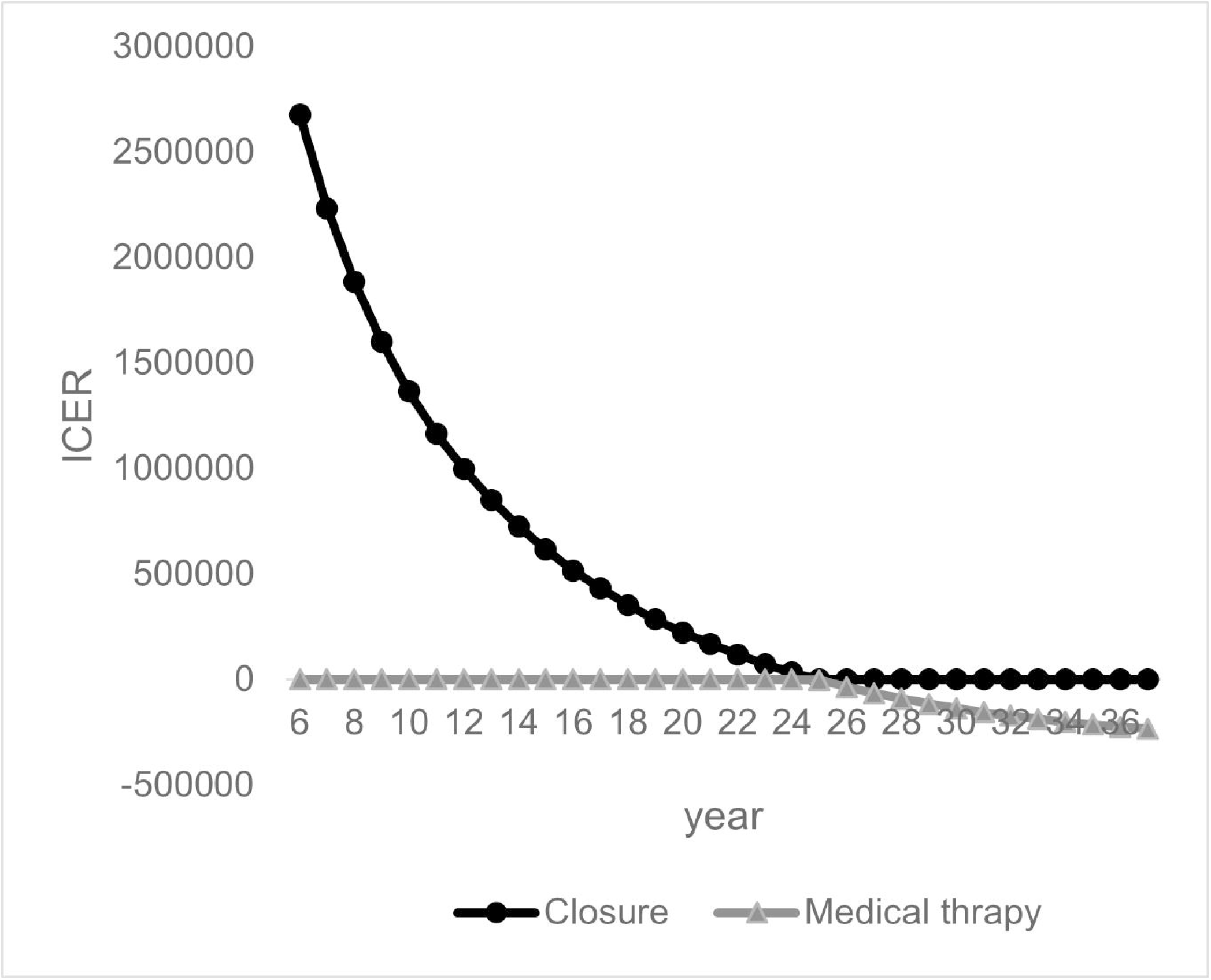
One-way sensitivity analysis: Time horizon closure vs. medical therapy. Time horizon closure vs. medical therapy, which is the second largest impact of parameter variation, is shown with ICER. Abbreviations. ICER, incremental cost-effectiveness ratio.

Probabilistic sensitivity analyses demonstrated that PFO closure was 50.3% cost-effective with a willingness-to-pay threshold of ¥5,000,000/QALY at a 5.9-year time horizon. (Figure 5) This result of this probabilistic sensitivity analysis means that PFO closure may be cost-effective, but there is still roughly a 50% chance of its not being cost-effective.

**Figure 5.**
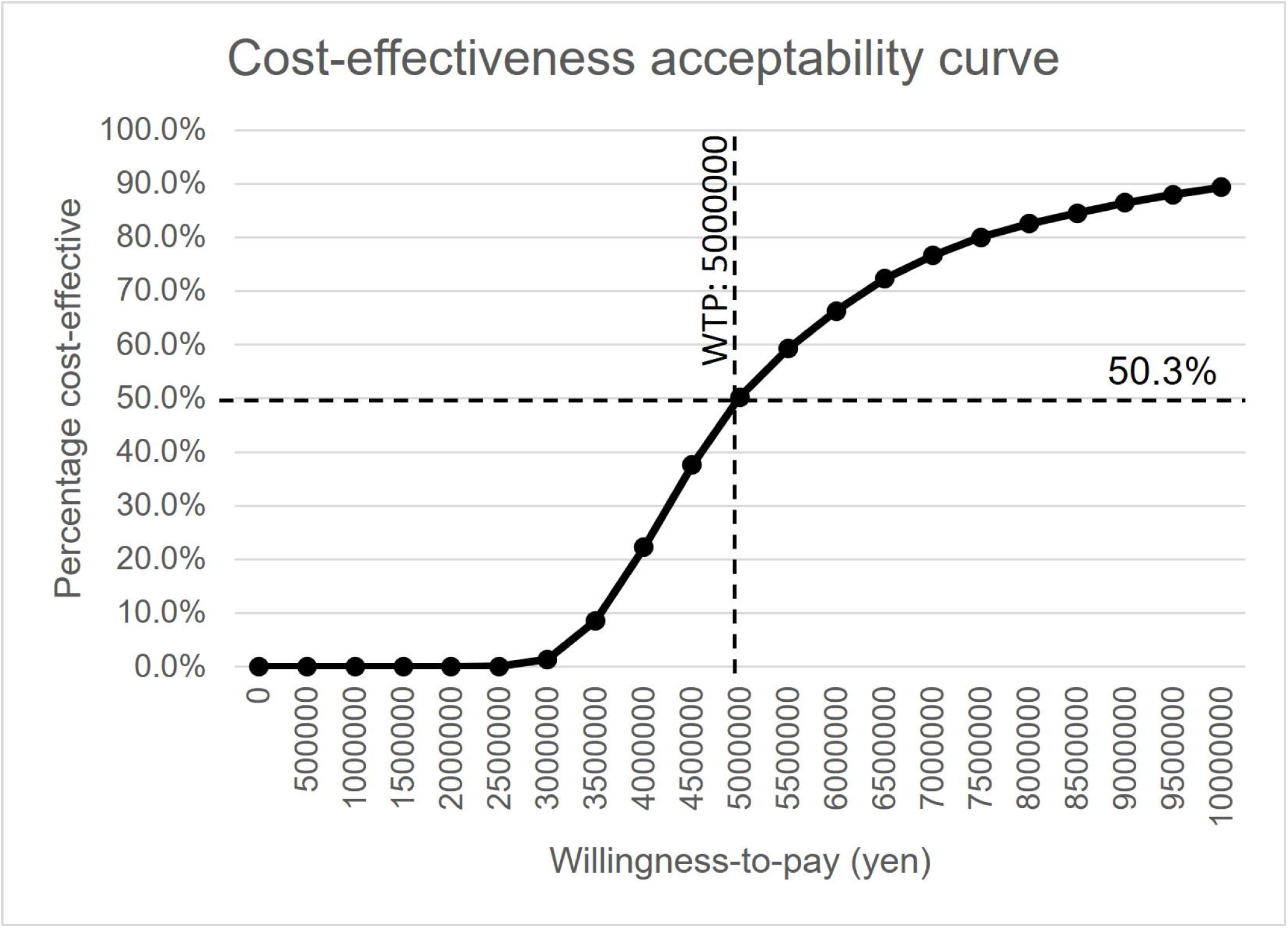
Cost-effectiveness acceptability curves. Willingness-to-pay (JP¥) is shown with percent cost-effectiveness. The dashed line is a willingness-to-pay threshold (WTP) of ¥5,000,000/QALY and the percent cost effectiveness is 50.3% at a WTP of ¥5,000,000/QALY. Abbreviations. WTP, Willingness-to-pay.

## Discussion

We evaluated the cost-effectiveness in Japan of PFO closure compared to medical therapy after cryptogenic stroke. The analysis revealed that PFO closure is cost-effective in the base case model, resulting in an ICER of ¥3,318,152 per QALY gained. Previous cost-effectiveness analyses outside Japan also revealed that PFO closure is cost-effective compared with medical therapy for cryptogenic stroke patients (11–16). A cost-effectiveness study in the U.S. (11) showed that PFO closure achieved an ICER of $21,049 at five years. In a U.K. study (15), ICER was reportedly £20,951 at four years. Both results are consistent with the Japanese situation. However, one-way sensitivity analysis showed that the stable state utility score difference between PFO closure and medical therapy had the largest impact on ICER. In other words, a stable state utility score difference below 0.051 rendered a result that was not cost-effective. This was the most important finding of this study because we assumed a base case utility score. Probabilistic sensitivity analysis also supports the result that PFO closure was only 50.3% cost-effective with a willingness-to-pay threshold of ¥5,000,000/QALY at a 5.9-year time horizon.

Assumptions of previous cost-effectiveness studies were based on expert opinions and some published literature, which indicated that PFO closure not only reduces recurrent stroke, but also patient anxiety about recurrent stroke. Study authors further indicated that PFO closure also reduces PFO-related migraine so that such reduction may improve the stable utility score, but we cannot be certain whether the utility score difference between PFO closure and medical therapy exceeds 0.051 without a reliable utility score for patients who have backgrounds similar to those in the RESPECT study. In addition, Japanese utility score evidence is important because of the difference in life expectancy and medical care systems in other countries. Hence, the Japanese stable utility score after PFO closure and medical therapy among patients with cryptogenic stroke and PFO aged ≤60 years needs to be determined through further studies. With new utility score evidence, Japanese criteria for PFO closure should be changed by releasing the quality indicator score to show that patient quality of life improves.

The goal of the basic act on stroke and cardiovascular disease countermeasures of 2018 is to promote healthy life expectancy. Our results may promote a cost-effective, healthy life expectancy by reducing recurrent ischemic strokes, which remain the leading cause of long-term disability. However, most reports on utility scores of stroke patients are from Europe and the United States, and very few studies with cases exist in Japan. When evaluating utility scores, it is necessary to use a reliable, validated measurement scale, but there are few such scales at present. Especially for new procedures or treatments, such as PFO closure, there is no evidence that shows the exact utility score after releasing RCTs all over the world. Without reliable utility scores, cost-effectiveness studies will always lack core information and can only offer hypotheses regarding economic impact.

In April 2019, the Ministry of Health Labour and Welfare (MHLW) of Japan introduced a cost-effectiveness evaluation system. The aim of the system is to utilize results of cost-effectiveness evaluations that will not be used for reimbursement decisions, but for post-listing price adjustments (28). Thus, utility score research should be performed using national grants for accurate evaluation of cost-effectiveness studies.

With an accurate utility score, PFO closure become a new component of a cardiovascular disease prevention strategy for the Japanese population. Our cost-effectiveness analysis of PFO closure supports this conclusion and can serve as a reference to consider whether political resources of the basic act on stroke and cardiovascular disease countermeasures should be allocated.

## Conclusion

From a healthcare payer perspective, PFO closure may be cost-effective compared with medical therapy in Japanese patients ≤60 years with cryptogenic strokes that are probably attributable to a PFO.

## Data Availability

All data are fully available without restriction and all relevant data are within the manuscript and its Supporting Information files.

## Author contributions

**Conceptualization:** Yoko Shijoh, Shota Saito, Zhehao Dai, Sachiko Ohde.

**Data curation:** Yoko Shijoh, Shota Saito, Zhehao Dai, Sachiko Ohde.

**Formal analysis:** Yoko Shijoh, Shota Saito, Zhehao Dai, Sachiko Ohde.

**Funding acquisition:** N/A

**Investigation:** Yoko Shijoh, Shota Saito, Zhehao Dai, Sachiko Ohde.

**Methodology:** Yoko Shijoh, Shota Saito, Zhehao Dai, Sachiko Ohde.

**Project administration:** Yoko Shijoh.

**Resources:** N/A

**Software:** N/A

**Supervision:** Sachiko Ohde.

**Validation:** Yoko Shijoh, Shota Saito.

**Visualization:** Yoko Shijoh, Shota Saito.

**Writing – original draft:** Yoko Shijoh.

**Writing – review & editing:** Yoko Shijoh, Shota Saito, Zhehao Dai, Sachiko Ohde.

## Acknowledgements

We wish to thank the timely help given by Dr. Ataru Igarashi, Associate Professor, Unit of Public Health and Preventive Medicine, School of Medicine, Yokohama City University, Yokohama, Japan, for useful discussions in analyzing data.

## Notes

### Competing Interest Statement

Yoko Shijoh is an employee of Bayer Yakuhin, Ltd., a pharmaceutical company located in Japan. This manuscript was a part of her dissertation during her obtaining MPH degree at Graduate School of Public Health St. Luke's International University. The Bayer Yakuhin, Ltd did not get involved in this work at all, neither financial support nor contents of study. All co-authors understand her situation and proceed our research purely academically.

### Funding Statement

The authors received no specific funding for this work.

### Author Declarations

All relevant ethical guidelines have been followed and the submission does not require an ethics statement.

